# Training the next generation of physician-scientists: a cohort-based program for MD-only residents and fellows

**DOI:** 10.1101/2022.12.19.22283532

**Authors:** Tina A. Solvik, Alexandra M. Schnoes, Thi A. Nguyen, Shannon L. Behrman, Elie Maksoud, Sarah S. Goodwin, Ethan J. Weiss, Arun Padmanabhan, David N. Cornfield

**Author notes:** **Corresponding Author** David N. Cornfield, Stanford University, 770 Welch Rd, Mail Code 5882, Stanford, California 94304. These authors contributed equally to this work (Tina A. Solvik, Alexandra M. Schnoes, Thi A. Nguyen).

## Abstract

**Importance:** Despite the importance of clinician-scientists in propelling biomedical advances, the proportion of physicians engaged in both hypothesis-driven research and clinical care continues to decline. Recently, multiple institutions have developed programs that promote MD-only physicians pursuing careers in science, but few reports on the impact of these are available.

**Objective:** To assess if a cohort-based training program for MD-only physician-scientists that includes didactic and experiential curricula favorably informs participants’ scientific development.

**Design:** The Chan Zuckerberg Biohub (CZB) Physician-Scientist Fellowship Program (PSFP) conducted a study from July 2020 to August 2023.

**Participants:** 24 inaugural program participants at UCSF and Stanford University (median postgraduate year at program start, 5.5; 17 clinical specialties represented; 10 [42%] identified as female; 7 [29%] identified as underrepresented in medicine).

**Exposures:** The CZB PSFP is a selective two-year career development program for MD-only physicians. Participants attended a two-week immersive training at the program outset, and subsequently, weekly curricular and scientific meetings throughout the program while conducting research.

**Main Outcomes and Measures:** Primary outcome measurements included pre-, 1-month, and 12-month assessments of confidence in research skills, career skills, and self-identification as scientists. Program satisfaction and feedback related to program curriculum and community were collected at 1 month, 6 months, and 12 months.

**Results:** After 12 months, 100% (N=16) reported satisfaction with the program and participants demonstrated increased confidence in research skills [median (IQR), 4.0 (2.5-5.0) pre-bootcamp to 5.5 (4.0-6.0) 12-mo], career skills significantly increased [median (IQR), 4.0 (4.0-5.0) pre-bootcamp to 5.5 (5.0-6.0) 12-mo], perceptions of belonging significantly increased [median (IQR), 4.0 (2.5-5.4) pre-bootcamp to 5.5 (5.1-7.0) 12-mo], and scientific identity significantly increased [median (IQR), 5.0 (4.0-5.5) pre-bootcamp to 6.0 (5.5-7.0) 12-mo].

**Conclusion and Relevance:** Participants demonstrated significant gains in confidence in core research and career skills as well as personal identification as scientists, demonstrating the efficacy of a longitudinal curriculum, peer support, and community building in fostering development as an investigator. The highly portable nature of this strategy may facilitate ready adoption and implementation at other institutions.

## Introduction

Over the past 100 years, physician-scientists have created important new knowledge across the entire spectrum of biomedical inquiry. Increasingly, successful development and application of novel therapies require the engagement of expert investigators from multiple disciplines.^1,2^ Creation of durable knowledge that enhances human health demands testing at the bedside and in the laboratory. Well-trained physician-scientists are uniquely equipped to pose scientific questions that link biomedical sciences to bedside care.^3^

Since the 1980s, the percentage of physicians dedicating significant professional time to research has declined from approximately 5% to 1.3%. Despite training more physicians, the absolute number of physician-scientists is declining while their average age is increasing.^4–7^ The relative percentage of research program grants awarded to MDs over age 50 increased from under 25% in 1977 to over 70% in 2012.^6,7^ Applying the power of fundamental scientific discovery to human health requires the number of early-career physician-scientists to be replenished and sustained.

Reports on the physician-scientist workforce have identified several major challenges including recruitment and retention, obtaining research funding, length and structure of training, limited mentor visibility, and tension between clinical and research responsibilities.^7,8^ To address these challenges, we built a novel program that provides substantive training, peer community, and mentorship to physicians interested in creating new knowledge focused on significant problems in human health, without significantly prolonging their training experience. The Chan Zuckerberg Biohub – San Francisco (CZ Biohub SF) Physician-Scientist Fellowship Program (PSFP) is a collaboration between the CZ Biohub SF, Stanford University, and the University of California, San Francisco (UCSF). The program targets MD-only physicians interested in undertaking hypothesis-driven, investigator-initiated scientific discovery while continuing to engage in clinical medicine.

The primary outcome goal of the CZB PSFP is to develop and support a pool of physician-scientists with fluency in the language of science and the ability to pose and answer biological questions to promote translation of basic science research into patient care. The program seeks to train and motivate participants to pursue a discovery-guided career, whether it is leading their own research group, acting as a clinical collaborator on research studies led by other scientists, or by being an engaged consumer of basic research studies that provides clinical care informed by new research findings. Importantly, the program seeks to serve as a model for academic institutions to adopt for facilitating physician-scientist development. As this program was only started in 2020, additional time will be required to effectively evaluate the long-term career outcomes of participants. Here, we present our initial evaluation of the CZB PSFP and its immediate impact on physician trainees’ development and identity as investigators.

## Methods

### Program Design

To address the barriers to MDs pursuing a physician-scientist career, we developed a structured two-year fellowship program that includes formal training to facilitate development of physicians as researchers. After considering the most critical determinants of long-term success for a physician-scientist career, the CZ Biohub SF PSFP emphasized three core factors: research and critical thinking skills (i.e., defining a research vision, identifying and addressing a gap in the field, experimental design, research resilience, and technical skills), career and professional development skills (i.e., strategic planning, identifying mentors, writing competitive grants, and designing and delivering high-quality research talks), and scientific identity (i.e., self-identification as a researcher, normalizing imposter fears, sense of belonging to scientific community).^9–11^ To achieve these goals, three primary methods were utilized: peer-to-peer mentorship and teaching to cultivate a community of physician-scientists at similar career stages, an intensive two-week introductory training in core scientific principles and tools, and a longitudinal curriculum focused on progressively building professional and research skills (**Figure 1**). The program provided financial support (50% of salary and benefits) to motivate dedicated research time, guidance related to mentorship and laboratory experiences, and coaching from invested senior, non-supervising physician-scientists. The program leadership includes two faculty co-directors from Stanford and UCSF, and, based at CZ Biohub SF, a program manager and a director of scientific programs. Consultants from the non-profit Science Communication Lab (SCL) designed the curriculum and facilitated its implementation in collaboration with the program leadership.

**Figure 1.**
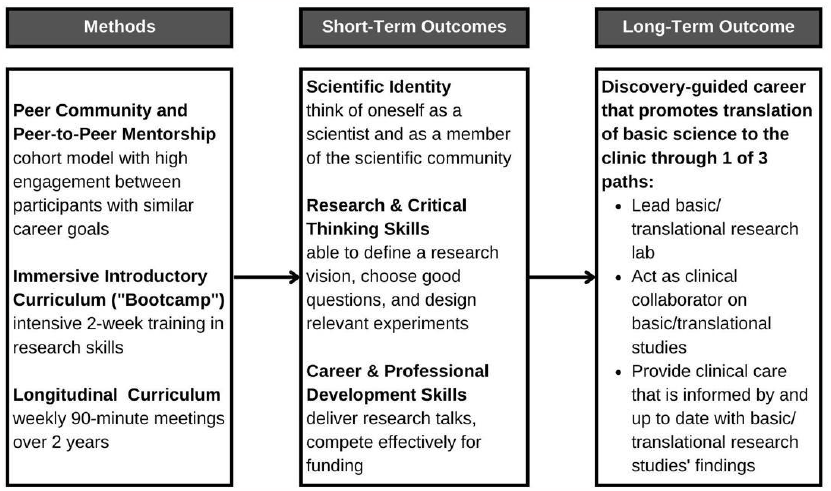
PSFP utilizes peer community and two curriculum methods to support the development of physician-scientists.

### Participants

Fellows held an MD, had not received nor were seeking a PhD, and were currently enrolled in or had completed an accredited post-graduate clinical training program at UCSF or Stanford. Participants committed to a 24-month program, with not less than 75% of their time dedicated to research, and maintained a minimal clinical effort of 20%.

### Curriculum and Delivery

The pillars of the PSFP curriculum were focused on competencies in research and critical thinking along with career and professional development (**Figure 2**). The competencies selected for this curriculum were based upon the experience of program leadership and staff and literature aligned with early-career researcher ^9–11^ and physician-scientist^12–14^ training programs. These competencies also address the specific areas of scientific mindset and belonging to the scientific community, as identified by CZ Biohub SF program directors. Supplemental figure eFigure 1 provides a comparison of the competencies identified in the literature for physician-scientists and the CZB PSFP. The curriculum included training courses produced by the Science Communication Lab, through their iBiology Courses initiative, designed to enhance career and professional development for researchers: (1) “Planning Your Scientific Journey,” focuses on research project development and planning;^15^ (2) “Let’s Experiment,” addresses experimental design;^16^ (3) “Share Your Research,” provides strategies to create and deliver an engaging research talk;^17^ (4) “Business Concepts for Life Scientists” teaches strategic business fundamentals for research and transition to independence.^18,19^ Lab-based training and didactic research training, including RNA sequencing, bioinformatics, computational microscopy, and CRISPR-based approaches were led by either CZ Biohub SF researchers or PSFP Fellows in a peer-taught manner.

**Figure 2.**
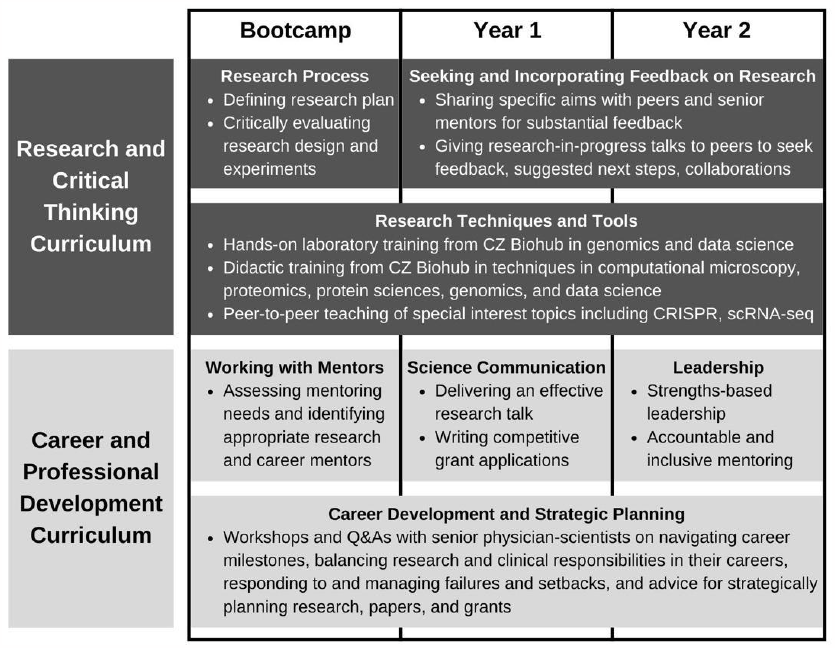
PSFP teaches key research and career skills throughout the curriculum.

At program outset each year in July, Fellows attended a group-based bootcamp consisting of nine two-hour trainings administered via a video conferencing platform in 2020 and 2021. In 2022, the training included six five-hour in-person sessions, including dedicated time for lab-based training at CZ Biohub SF headquarters. From August through June each year, participants attended weekly 90-minute sessions; in 2020-2021, sessions were virtual only, and in 2022, sessions were a mixture of virtual and in-person to promote further community building. In preparation for group meetings, Fellows completed pre-session work, including reviewing videos and completing writing assignments. Synchronous sessions included didactic teaching, facilitated group discussion, and application of the skills in breakout rooms.

### Procedures

We examined learning outcomes and program satisfaction using pre- and post-course surveys. Assessments used a 7-point Likert scale from “not at all confident” to “very confident” for learning outcomes and “extremely dissatisfied” to “extremely satisfied” for program satisfaction. The survey questions to assess confidence in skill development were based on the competencies identified by PSFP program directors and program staff (eFigure 1). The survey questions to assess the concepts of self-efficacy and self-identity as a scientist were adapted from Estrada et al. (2011).^9^

### Statistical Analysis

Pre- and post- results after 1-month and 12-month in the program (post-bootcamp) were compared using nonparametric, matched-pairs rank tests (Wilcoxon). If both pre- and post- surveys were completed by the same participant at either timepoint, the data were included in the analysis. We chose to use non-parametric analyses in this study due to the non-normal distribution of the data. We used GraphPad statistical software, version 9, to analyze data. Survey data to compare different competencies (eTable1) were also analyzed descriptively. Our sample of participants was a selection sample based on the application process, and therefore, not necessarily representative of other MD-only physician-scientist training programs.

## Results

### Program Participants

The program sought participant diversity in gender, representation, postgraduate year (PGY), and clinical specialty (**Table 1**). Of 24 Fellows in the first three cohorts of the program (2020-2022), 10 (41.7%) were female and 7 (29.2%) self-identified as underrepresented in medicine according to the AAMC definition.^20^ The average PGY for Fellows at the start of the program was 5.8 (SD 1.4). The most common clinical specialties included cardiology, hematology/oncology, infectious disease, pulmonary and critical care medicine, and surgical subspecialties.

**Table 1.** Characteristics of participants to date, 2020-2023.

| Characteristic | Participant No. (%) |
| --- | --- |
| Total participants | 24 (100.0) |
| Total applicants | 76 (316.7) |
| Gender |  |
| Female | 10 (41.7) |
| Male | 14 (58.3) |
| Representation in medicine |  |
| Underrepresented | 7 (29.2) |
| Well-Represented | 15 (62.5) |
| Prefer not to answer | 2 (8.3) |
| Institution |  |
| Stanford University | 10 (41.7) |
| UCSF | 14 (58.3) |
| PGY at program start |  |
| Mean (SD) | 5.8 (1.4) |
| 3 | 1 (4.2) |
| 4 | 2 (8.3) |
| 5 | 8 (33.3) |
| 6 | 7 (29.2) |
| 7 | 2 (8.3) |
| 8 | 3 (12.5) |
| 9 | 1 (4.2) |
| Clinical Specialty |  |
| Allergy and Immunology | 1 (4.2) |
| Cardiology (Pediatric, Adult) | 1 (4.2), 2 (8.3) |
| Clinical Pathology/Laboratory Medicine | 1 (4.2) |
| Dermatology | 1 (4.2) |
| Gastroenterology | 1 (4.2) |
| Hematology/Oncology (Pediatric, Adult) | 4 (16.7), 2 (8.3) |
| Infectious Disease (Pediatric, Adult) | 1 (4.2), 1 (4.2) |
| Neurology | 1 (4.2) |
| Pulmonary and Critical Care Medicine (Pediatric, Adult) | 2 (8.3), 2 (8.3) |
| Rheumatology | 1 (4.2) |
| Surgery |  |
| General | 1 (4.2) |
| Neurological | 1 (4.2) |
| Vascular | 1 (4.2) |

### Confidence in Skills Related to Research and Career and Professional Development

Fellows reported an increase in confidence in the broad categories of research skills and career skills at one month (post-bootcamp) (**Figure 3**). The median confidence in research skills increased from 4.0 (IQR= 2.5-5.0) pre-bootcamp to 5.0 (IQR=4.0-5.5) post-bootcamp. Median confidence in career skills also increased from 4.0 (IQR= 3.0-5.5) pre-bootcamp to 5.0 (IQR=4.0-5.5) post-bootcamp. Fellows also reported increased perceptions of belonging with a median of 4.0 (IQR= 2.5-5.5) pre-bootcamp to 5.0 (IQR=5.0-5.5) post-bootcamp. There was no increase in perceptions of scientific identity after 1-month with a median 5.0 (IQR= 4.0-5.5) pre-bootcamp and 5.0 (IQR=5.0-6.0) post-bootcamp.

**Figure 3.**
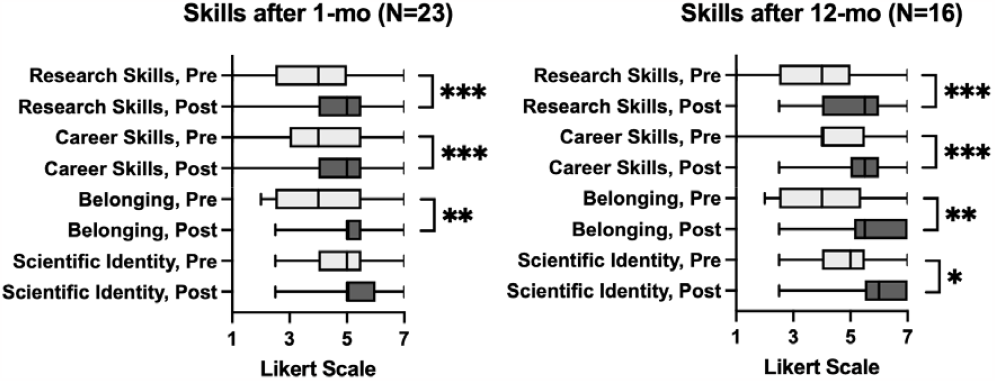
Participants report increased confidence in research skills, career and professional development skills, and research identity after 1-month and 1-year in the program. Learning Outcomes for Immersive, 1-mo and 12-mo training include all three cohorts (2020-22). Results represent matched-pair pre-/post-survey responses (1-mo, N=23; 12-mo, N=16). Pre-survey is given in July when each cohort starts. For research and career skills, responses could range from 1 (not at all confident) to 7 (very confident). For research identity and belonging, responses could range from 1 (strongly disagree) to 7 (strongly agree). Boxes represent median and interquartile range for paired data; bars represent min and max values. Statistical significance for nonparametric Wilcoxon matched-pairs rank tests, \**p* ≤ 0.05, \*\**p* ≤ 0.01, \*\*\**p* ≤ 0.001

The increases in skill development persisted at 12 months. Confidence in research skills significantly increased [median (IQR), 4.0 (2.5-5.0) pre-bootcamp to 5.5 (4.0-6.0) 12-mo], career skills significantly increased [median (IQR), 4.0 (4.0-5.0) pre-bootcamp to 5.5 (5.0-6.0) 12-mo], perceptions of belonging significantly increased [median (IQR), 4.0 (2.5-5.4) pre-bootcamp to 5.5 (5.1-7.0) 12-mo], and Scientific Identity significantly increased [median (IQR), 5.0 (4.0-5.5) pre-bootcamp to 6.0 (5.5-7.0) 12- mo].

The research skills category constituted eight distinct elements including identifying experimental bias, choosing a good research question, and acquiring new experimental methods. Relative to levels prior to matriculation in the program, median confidence in research skill development was higher and interquartile ranges (IQR) did not overlap in all eight elements at 12 months for unpaired survey results (**eTable 1**). The career skills comprised six specific elements. Four areas had the biggest areas of growth at 12 months (developing a strategic plan for research, defining strategic goals for career, writing successful grants, delivering high quality research talks), and higher median confidence with non-overlapping IQR ranges (**eTable 1**). It was not surprising to see that confidence was relatively unchanged in the competency of writing papers as that skill was not explicitly addressed in the curriculum. The category of identifying and recruiting diverse mentors had relatively unchanged confidence levels at 12-months, which might reflect sound mentorship choices even prior to joining the program.

### Self-perception as a Scientist

The community building efforts and training positively impacted Fellows’ perceptions of belonging to the scientific community and self-identification as a scientist (referred in the figure as “scientific identity”, **Figure 3**). After one month of the program, post-bootcamp, the Fellows agreed with the statements “I have a strong sense of belonging to the community of scientists” and “I have come to think of myself as a researcher.” At 1-mo a significant increase was only reported for perceptions of belonging. At 12-months there was a significant increase in both perceptions of belonging to the scientific community and scientific identity.

### Satisfaction with the Program

Program satisfaction was present at all timepoints measured and no Fellow reported being dissatisfied (**Table 2**). In response to open-ended questions about program satisfaction and perceived value added, participants emphasized the community and curriculum as program highlights, saying:

**Table 2.** Satisfaction with program after 1 month, 6 months, and 12 months in program. Participants include 2020, 2021, and 2022 cohorts. Pre survey taken in July when each cohort started. 7-point Likert scale, from “extremely satisfied (7)” to “extremely dissatisfied (1).” Satisfied is 5 and above.

| TIME IN PROGRAM | MEAN (SD) | N Satisfied (%) |
| --- | --- | --- |
| 1 month, after the SUMMER intensive training (N=21) | 6.3 (0.64) | 21 (100) |
| 6 months, after weekly FALL training (N=18) | 6.1 (0.85) | 18 (100) |
| 12 months, after weekly SPRING training (N=16) | 6.0 (0.88) | 16 (100) |

> *“I really appreciate the sense of community with the fellows program. Outside of this program, I would have very limited exposure to other MD-only physician-scientists in training, so it is great to be part of this community!”*
>
> *“The structure, community, and consistency of the program have been very useful over the past year. The talks have been poignant, on topic, and tailored to our specific needs as young career physician scientists*.*”*
>
> *“The camaraderie was the best part. Hearing the stories from everyone normalized my own feelings, and the community atmosphere was really special*.*”*
>
> *“I know that I will use the skills I learned from [the program] in my future academic career – including research planning, design, and implementation, and also presentation skills, critical thinking, and career planning*.*” (email communication from a 2020 cohort participant, shared with written permission)*

## Discussion

Despite the need for physicians capable of asking and answering scientific questions with clinical significance and contributing meaningfully to team science, relatively fewer physicians are pursuing a discovery-intensive career path. The challenges to a physician-scientist career are myriad and well documented including: (1) the time required to gain scientific competence, (2) the perceived need to choose either science or medicine, (3) the paucity of similarly inclined peers, and (4) the absence of role models.^21^ Our program sought to explicitly address each of these challenges.

First, the two-year CZ Biohub SF PSFP was designed to maximize scientific skills, without significantly prolonging the training experience of physician-scientists. To build skills, confidence, and teach essential research methods, the program included an immersive introductory two-week curriculum in July of the matriculating year and a longitudinal curriculum provided in weekly seminars from August through June for the next two years. The present report demonstrates that Fellows found value, and even identity in the curriculum, with progressive improvement in each of the research and critical thinking-related metrics assessed. The effects on career and skill development were similarly pronounced, progressive, and durable.

Second, the program emphasized the feasibility of a dual career in science and medicine. Administratively, this was promoted through the requirement for ongoing engagement in clinical medicine for no less than 20% effort, demonstrating the potential to be highly competent and engaged as both a clinician and scientist and the synergy between the domains. The curriculum was designed to build skills, confidence, and exposure to research. Arguably, the Fellows’ strong agreement with the statement “I have come to think of myself as a researcher” represents the most compelling data in support of the program. Self-identification as a scientist has been found to be a significant predictor of a trainee’s persistence in science.^22–27^

Third, the program focused on fostering community and providing peer-to-peer and near-peer interactions to normalize imposter fears and bolster the determination to pursue both science and medicine. As a cohort, participants were encouraged to discuss their pursuit of a discovery-focused career and build self-confidence by giving research presentations and peer teaching technical concepts. The findings in this report suggest that these efforts were successful, as the sense of belonging to a community of scientists increased markedly after the two-week bootcamp alone. Further evidence of the success of the community creation includes narrative comments from participants, with a high value placed on belonging to a community. By including trainees across clinical specialties, the program created a new peer and near-peer support community.

Finally, to address the issue of mentoring and role modeling, the program was designed to provide exposure to clinicians at multiple career stages. The co-directors are each engaged clinicians directing NIH-funded, discovery-based research programs and attended over 90% of the cohort meetings. Furthermore, early-, mid-, and late-career physician-scientists were engaged in candidate interviews, career discussion panels and networking lunches during the bootcamp, and career seminars throughout the academic year.

The program was specifically designed to be easily portable, thus promoting creation of similar programs at institutions interested in advancing physician-scientist training. By identifying the critical determinants of formation of a researcher identity for MD-only investigators, institutions interested in increasing the supply of physicians-scientists can more expeditiously and efficiently deploy resources to craft effective training programs with a high likelihood of success.

## Conclusions

The early results of this novel physician-scientist training program suggest that concentrating on creating a cohort of aspiring MD-trained scientists and providing focused curriculum over two years can foster the development of scientific identity and skill acquisition. The program targeted participants at an especially vulnerable point in training. In conclusion, iterative, structured scientific training paired with intentional community building can increase the competence, commitment, and confidence of physician-scientists engaged in discovery to pursue a research career. Whether the successes of the present program can be translated beyond UCSF and Stanford remains unknown, though the program’s success with a variety of physician-scientists representing a wide array of medical and surgical disciplines argues for the viability and broad applicability of the present strategy.

## Data Availability

All data produced in the present study are available upon reasonable request to the authors.

## Acknowledgments

We thank Bill Burkholder, PhD [CZ Biohub SF], Elliot Kirschner [Science Communication Lab], Catherine Tan [CZ Biohub SF], Roger Nys, MPA [CZ Biohub SF], and Lubert Stryer, MD [CZ Biohub SF, Stanford University] for their support and contributions to the program. Roger Nys received financial compensation for his administrative support of the program. We thank Amanda Butz, PhD, Vivekinan Ashok, PhD, and Rebekah Layton, PhD for consultation about statistical analysis. We also thank our colleagues at Stanford University, UCSF, UC Berkeley, and CZ Biohub SF who have provided their time and support as speakers, panelists, mentors, advisors, reviewers, and interviewers, and the administrative support for the program provided by our university partners.

## Supplementary Material

**Supplemental Table 1.** Increased Confidence in Skills after 12-months in program. Descriptive statistics for unpaired survey responses. Reported are median and interquartile range (IQR) for data pre- and post-time points, as well as mean and standard deviation (SD). Participants include 2020, 2021, and 2022 cohorts during the first year of the program. Pre-survey taken in July when each cohort started. Post-survey taken in June the following year (POST-12mo). For Research and Career Skills, responses could range from 1 (not at all confident) to 7 (very confident). For Scientific Identity and Belonging, responses could range from 1 (strongly disagree) to 7 (strongly agree).

| TOPIC | PRE,<br>N=23<br>median<br>(IQR) | POST 12-mo, N=16<br>median<br>(IQR) |  | PRE,<br>N=23<br>mean<br>(SD) | POST<br>12-mo,<br>N=16<br>mean<br>(SD) |
| --- | --- | --- | --- | --- | --- |
| **RESEARCH - Research Critical Thinking, Experimental Design & Methods, N=23** |  |  |  |  |  |
| Defining my research vision | 4.0 (3.0-5.5) | 5.5 (5.5-6.0) |  | 4.46 (1.15) | 4.42 (1.30) |
| Describing how my research will fill a gap in the field | 5.0 (4.0-5.5) | 5.5 (5.5-6.0) |  | 4.76 (1.27) | 4.54 (1.10) |
| Choosing a good research question | 4.0 (3.0-4.0) | 5.5 (5.5-5.9) |  | 4.20 (1.12) | 4.69 (0.98) |
| Designing experiments that will provide interpretable results | 3.0 (2.5-4.0) | 5.5 (4.2-5.9) |  | 3.52 (0.98) | 4.19 (1.42) |
| Acquiring new experimental methods | 3.0 (2.5-4.0) | 4.7 (4.0-5.9) |  | 3.37 (1.27) | 4.74 (1.23) |
| Choosing appropriate model systems for my research | 3.0 (2.5-4.0) | 5.5 (4.0-6.0) |  | 3.34 (1.22) | 4.83 (1.17) |
| Identifying experimental bias | 3.0 (2.0-4.0) | 5.5 (4.0-6.0) |  | 3.52 (1.35) | 4.96 (1.16) |
| Discussing the impact of racial bias in research | 3.0 (2.5-4.0) | 5.0 (4.0-5.5) |  | 3.69 (1.63) | 5.09 (1.33) |
| **CAREER - Career and Professional Development & Goals, N=23** |  |  |  |  |  |
| Developing a strategic plan for my research | 4.0 (2.5-4.0) | 5.5 (5.0-5.5) |  | 3.85 (1.10) | 4.91 (1.22) |
| Defining strategic career goals for myself | 4.0 (2.5-4.0) | 5.5 (5.5-6.0) |  | 4.02 (1.17) | 5.15 (1.10) |
| Identifying and recruiting diverse mentors | 4.0 (3.0-5.0) | 5.5 (4.2-6.0) |  | 4.39 (1.11) | 5.28 (1.36) |
| Writing grants that could be successfully funded | 3.0 (2.5-4.0) | 5.2 (4.0-5.5) |  | 3.19 (1.04) | 4.26 (1.10) |
| Designing and delivering high quality research talks | 4.0 (3.0-5.5) | 5.5 (5.5-6.0) |  | 4.54 (1.30) | 4.76 (1.23) |
| Writing papers that could be accepted for publication | 4.0 (3.0-5.5) | 5.5 (4.2-5.5) |  | 4.50 (1.48) | 4.71 (1.35) |
| **You and Your Research Identity, N=23** |  |  |  |  |  |
| I have a strong sense of belonging to the community of scientists. | 3.0 (2.5-4.0) | 5.5 (5.1-7.0) |  | 3.98 (1.51) | 5.13 (1.13) |
| I have come to think of myself as a researcher. | 5.0 (3.0-5.5) | 6.0 (5.5-7.0) |  | 4.85 (1.42) | 5.26 (1.13) |

**Supplemental Figure 1.**
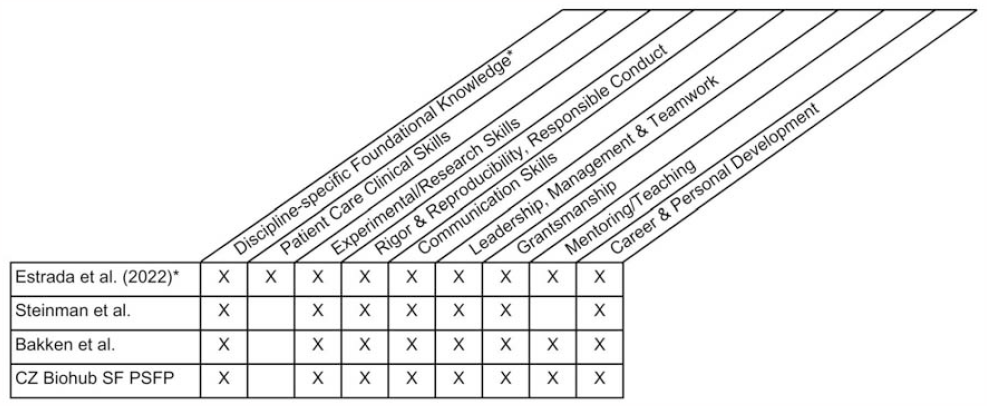
Comparison of CZB PSFP core competencies to three published physician-scientist competencies. The main competencies for the CZ Biohub SF PSFP are compared to lists of published competencies ^12–14^ for physician-scientist training. The CZ Biohub SF PSFP includes training for each of the competencies outlined in this literature, except for patient care clinical skills. Clinical care skills, though critically important for the physician-scientist, were addressed in each fellow’s clinical hours and not in the curriculum of this program. *The comparison is organized using the categories defined in Estrada et al. (2022)^12^ as those competencies are comprehensive and concise, and are most similar to how we have conceptualized our own program competencies.

## Notes

### Competing Interest Statement

Arun Padmanabhan serves on the Board of Directors of the Sarnoff Cardiovascular Research Foundation. David N. Cornfield serves on the Physician-Scientist Advisory Board of the Burroughs Wellcome Fund.

### Funding Statement

This study did not receive any funding.

### Author Declarations

Ethics committee/IRB of University of California, San Francisco gave ethical approval for this work (IRB 15-17981).

### Summary of Updates

The manuscript has been revised to include updated statistical analysis, a new figure comparing the program competencies to 3 other published lists of physician-scientist competencies, and an updated discussion.

